# Efficacy and Safety of neoadjuvant stereotactic body radiotherapy plus adebrelimab and chemotherapy for triple-negative breast cancer: A pilot study

**DOI:** 10.1101/2023.08.14.23294091

**Authors:** Guanglei Chen, Xi Gu, Xu Zhang, Xiaopeng Yu, Yu Zhang, Jinqi Xue, Ailin Li, Yi Zhao, Guijin He, Meiyue Tang, Fei Xing, Jianqiao Yin, Xiaobo Bian, Ye Han, Shuo Cao, Chao Liu, Xiaofan Jiang, Keliang Zhang, Yan Xia, Huajun Li, Nan Niu, Caigang Liu

## Abstract

**Background:** Emerging data have supported the immunostimulatory role of radiotherapy, which could exert a synergistic effect with immune checkpoint inhibitors (ICIs). With proven effective but suboptimal efficacy of ICI and chemotherapy in triple-negative breast cancer (TNBC), we designed a pilot study to explore the efficacy and safety of neoadjuvant stereotactic body radiotherapy (SBRT) plus adebrelimab and chemotherapy in TNBC patients.

**Methods:** Treatment-naïve TNBC patients received two cycles of intravenous adebrelimab (20mg/kg, every 3 weeks), and SBRT (24Gy/3f, every other day) started at the second cycle, then followed by six cycles of adebrelimab plus nab-paclitaxel (125 mg/m^2^ on days 1 and 8) and carboplatin (area under the curve 6 mg/mL per min on day 1) every 3 weeks. The surgery was performed within 3-5 weeks after the end of neoadjuvant therapy. Primary endpoint was pathological complete response (pCR, ypT0/is ypN0). Secondary endpoints included objective response rate (ORR), residual cancer burden (RCB) 0-I and safety.

**Results:** 13 patients were enrolled and received at least one dose of therapy. 10 (76.9%) patients completed SBRT and were included in efficacy analysis. 90% (9/10) of patients achieved pCR, both RCB 0-I and ORR reached 100% with 3 patients achieved complete remission. Adverse events (AEs) all-grade and grade 3-4 occurred in 92.3% and 53.8%, respectively. 1 (7.7%) patient had treatment-related serious AEs. No radiation-related dermatitis or death occurred.

**Conclusions:** Adding SBRT to adebrelimab and neoadjuvant chemotherapy led to a substantial proportion of pCR with acceptable toxicities, supporting further exploration of this combination in TNBC patients.

**Funding:** This research did not receive any specific grant from funding agencies in the public, commercial, or not-for-profit sectors.

**Clinical trial number:** NCT05132790.

## Introduction

Triple-negative breast cancer (TNBC), defined as estrogen receptor, progesterone receptor and human epidermal growth factor receptor 2 (HER2) negative, accounts for approximately 10-20% of all breast cancers(Costa et al., 2017). Characterized by higher tumor mutation burden and more intensive tumor-infiltrating lymphocytes (TILs) in tumor microenvironment (TME)(Loi et al., 2019), TNBC is more sensitive to immune checkpoint inhibitors (ICIs) than any other subtypes(Keenan & Tolaney, 2020). Nowadays, the combination with pembrolizumab (a PD-1 blockade) and standard chemotherapy is recommended for TNBC patients with stage II-III according to National Comprehensive Cancer Network (NCCN) guidelines(Gradishar et al., 2022). However, there is still around 40% of TNBC patients cannot achieve a pathological complete response (pCR) according to KEYNOTE-522 (Schmid et al., 2020) and IMpassion031 (Mittendorf et al., 2020) trials, which calls for more promising strategies.

Radiotherapy (RT) is the most critical locoregional treatment in solid malignancies and approximately half of patients may receive RT during the entire treatment period (Atun et al., 2015; Citrin, 2017). Stereotactic body radiotherapy (SBRT), a novel technique with higher doses of radiation delivery to the tumor lesion in a smaller number of fractions, can shorten treatment dduration and reduce exposure to the surrounding tissues (Chen et al., 2020). SBRT has already been widely applied to advanced breast cancer as salvage treatment targeting to osseous and brain metastasis or other oligometastatic sites (Nicosia et al., 2022; Viani et al., 2021). Preoperative SBRT may be advantageous for downstaging the tumor to enable breast conservation and improving pCR rate (Piras et al., 2023), with multiple trials investigating neoadjuvant SBRT in various malignancies (Holyoake et al., 2021; Kishi et al., 2020; Liu et al., 2022; Novikov et al., 2021) are currently ongoing. In addition, preclinical evidence suggested that SBRT has immunomodulatory properties by inducing cancer cells necrosis and releasing neoantigens to boost neoantigen–specific immune response and upregulate the expression of PD-L1 on tumor cells (Deng et al., 2014; Han et al., 2022; Park et al., 2015). Given these, it would be promising to combine SBRT with ICIs and chemotherapy under the neoadjuvant setting.

Therefore, we conducted a prospective pilot study to determine the efficacy and safety of neoadjuvant SBRT in combination with adebrelimab (SHR-1316), a potent selective PD-L1 inhibitor, plus nab-paclitaxel and carboplatin in patients with newly-diagnosed early or locally advanced TNBC, to validate the feasibility of this novel regimen in neoadjuvant treatment of TNBC.

## Methods

### Patients

Previously untreated patients aged between 18 and 75 years, with histologically confirmed invasive triple-negative breast cancer (defined as negative estrogen receptor, progesterone receptor and HER2 status by American Society of Clinical Oncology/College of American Pathologists guidelines) and pathological tumor size ≥ 2.0 cm in MRI assessment, were eligible for enrollment. Other inclusion criteria included Eastern Cooperative Oncology Group performance score of 0-1, adequate marrow, hepatic, renal and cardiac function.

Key exclusion criteria included bilateral, inflammatory or occult breast cancer; active or a history of autoimmune disease; use of glucocorticoids or other immunosuppressive therapy within 2 weeks before the first study dose; a history of interstitial pneumonia; active tuberculosis; pregnancy, lactation, and refusal to use contraception.

### Study design and treatment

This single-arm, prospective study was approved by the Institutional Review Board of Shengjing Hospital, China Medical University and performed in accordance with the Declaration of Helsinki and Good Clinical Practice. Written informed consent was obtained from each patient. This study was reported following the Strengthening the Reporting of Observational Studies in Epidemiology (STROBE) reporting guidelines.

All patients intravenously received 8 cycles of adebrelimab (20 mg/kg every 3 weeks). SBRT (24Gy/3f) targeted to breast lesion started at the second cycle every other day, and 6 cycles of nab-paclitaxel (125 mg/m^2^ on days 1 and 8) and carboplatin (area under the curve 6 mg/mL per min on day 1) was given every 3 weeks since the third cycle.

Patients who completed or discontinued the neoadjuvant treatment could recieve surgery. If disease progressed during the neoadjuvant phase, the patient would either proceed to surgery or receive alternative neoadjuvant therapy. Surgery was performed 3-5 weeks after the last dose of neoadjuvant therapy. Recommended surgery and adjuvant therapy were administered as per local guidelines or institutional standards.

### Outcomes

The primary endpoint was pCR rate in the breast and axillary lymph nodes (ypT0/is ypN0). Secondary endpoints included ORR before surgery (defined as the proportion of patients with complete or partial response according to the Response Evaluation Criteria in Solid Tumors [RECIST] version 1.1), the proportion of residual cancer burden (RCB) 0-I, and safety according to the Common Terminology Criteria for Adverse Events (CTCAE) version 5.0.

### Statistical analysis

Efficacy was assessed in the modified intention-to-treat population, which included patients who had undergone radiotherapy. Safety was evaluated in all recruited patients who received at least one dose of study drug. All statistical analyses were conducted using SAS 9.4 (North Carolina, USA). Continuous data are presented as mean and standard deviation (SD), or mean and 95% confidence interval (CI). Categorical data are expressed as frequency and percentage. The 95% CIs of pathological complete response rate, proportion of patients with RCB 0-I, and ORR were estimated using the Clopper-Pearson method.

## Results

### Patient characteristics

Between December 2021 and January 2023, 13 patients were enrolled in the trial (***Figure 1***). The baseline characteristics are shown in ***Table 1***. The median age was 51 years (range 31-68) and the median tumor size was 33mm (range, 22-72). Lymph node involvement was seen in 53.8% (7/13) patients and 46.2% (6/13) patients had stage III breast cancer at baseline.

**Table 1.** Baseline characteristics.

| **Characteristic** | **Patients (n=13)** |
| --- | --- |
| **Age (median, range)** | 51 (31-68) |
| **Age Group, years** |  |
| ≤ 50 | 6 (46.2%) |
| > 50 | 7 (53.8%) |
| **Menopausal status** |  |
| Premenopausal | 6 (46.2%) |
| Postmenopausal | 7 (53.8%) |
| **Tumor size** |  |
| T2 | 10 (76.9%) |
| T3 | 3 (23.1%) |
| **Lymph node status** |  |
| N0 | 6 (46.2%) |
| N1 | 2 (15.4%) |
| N2 | 5 (38.5%) |
| **Clinical stage** |  |
| IIA | 6 (46.2%) |
| IIB | 1 (7.7%) |
| IIIA | 6 (46.2%) |
| **Tumor grade** |  |
| II | 6 (46.2%) |
| III | 5 (38.5%) |
| Unknown | 2 (15.4%) |
| **HER2 expression** |  |
| Negative | 8 (61.5%) |
| 1+ | 3 (23.1%) |
| 2+, FISH- | 2 (15.4%) |

**Figure 1.**
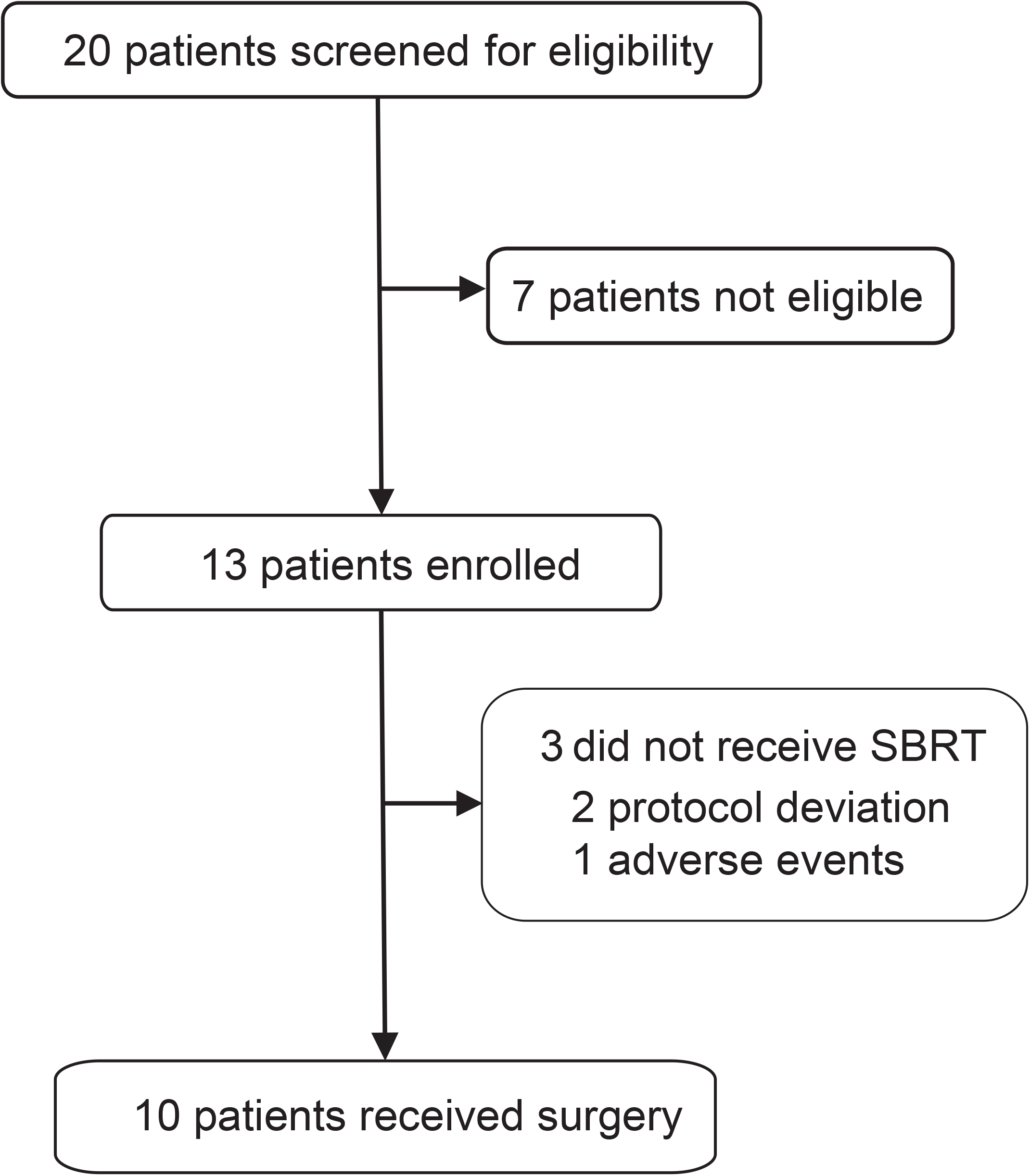
Flowchart of the trial. SBRT, stereotactic body radiotherapy.

### Outcomes

Among the 13 treated patients, 2 (15.4%) were excluded after the first dose of adebrelimab due to protocol deviation and 1 (7.7%) discontinued after the second dose of adebrelimab due to adverse event, thus only 10 (76.9%) who underwent neoadjuvant SBRT and surgery (the modified intention-to-treat population) were available for efficacy evaluation (***Figure 1***). 9 of the ten efficacy-evaluable patients (90%, 95% CI 55.5%-99.8%) achieved pCR in the breast and axillary lymph nodes, and the rates of RCB 0-I was 100% (95% CI 67.9%-100.0%). The radiologic ORR was 100% (95% CI 67.9%-100.0%, ***Table 2* and *Figure 2***) with 3 patients achieved complete radiographic response. 4 patients with positive lymph node at baseline had nodal downstaging to N0 after neoadjuvant treatment.

**Table 2.** Pathological and clinical response.

| Variable | Patients (n=10) |
| --- | --- |
| **Total pathological complete response** | 9 (90%) |
| **Residual cancer burden score** |  |
| 0 | 9 (90%) |
| I | 1 (10%) |
| II | 0 |
| III | 0 |
| **Radiological response** |  |
| Complete response | 3 (30%) |
| Partial response | 7 (70%) |
| Stable disease | 0 |
| Objective response rate | 10 (100%) |

**Figure 2.**
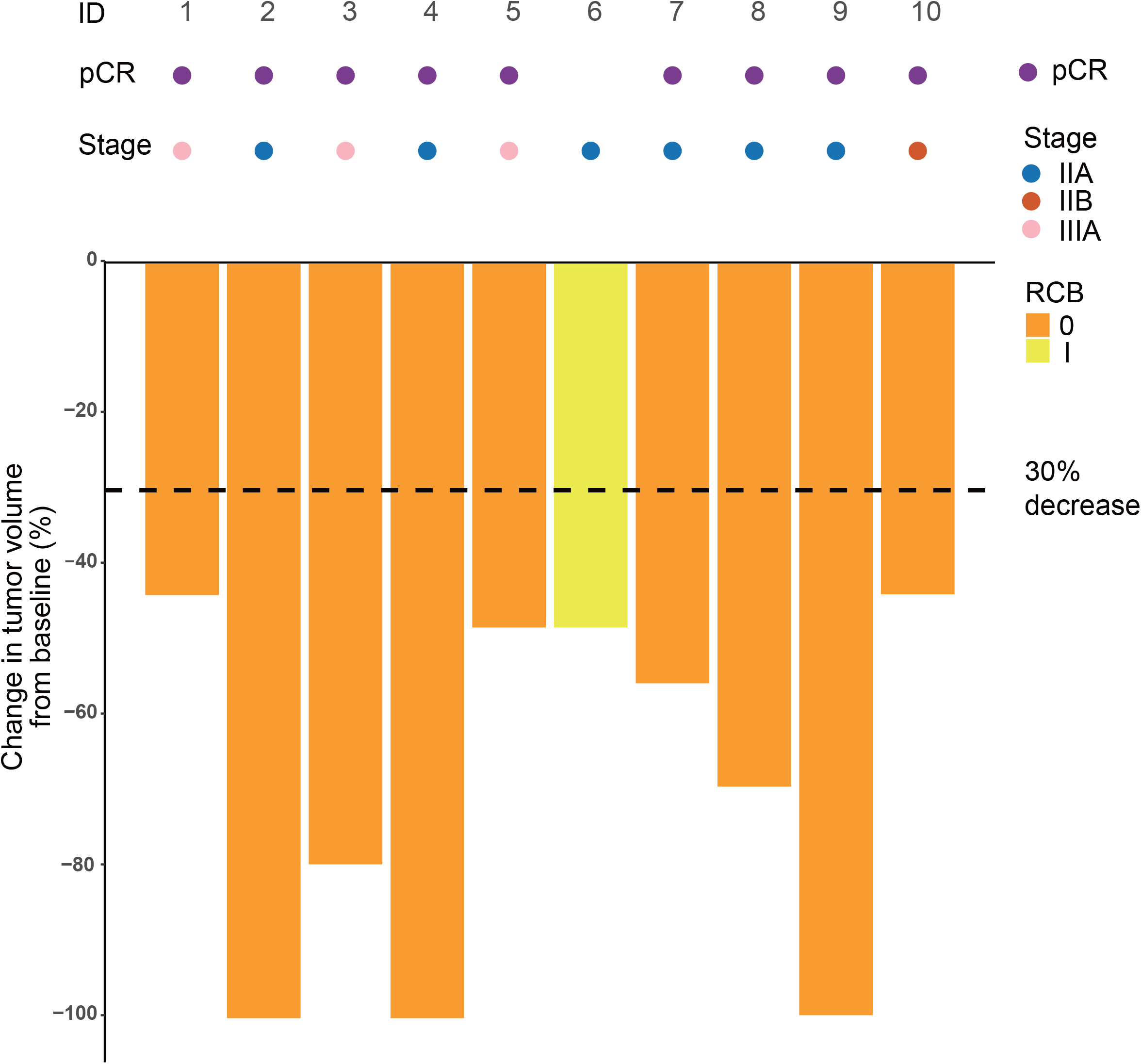
Swimming plot which demonstrate the pCR, RCB and radiological response profile of ten modified intention-to-treat population who received radiotherapy and undergone surgery. Each round dot or column indicates a patient. Colors indicate different clinical stage. ID, identity; pCR, pathological complete response; RCB, residual cancer burden.

### Safety

AEs were reported in all 13 patients (***Table 3***). The incidence of any grade AEs was 92.4%. The grade 3 or higher treatment-related adverse events occurred in 53.9% of the patients, including neutropenia (30.8%), anemia (7.7%), leukopenia (7.7%), thrombocytopenia (7.7%), creatine phosphokinase elevation (7.7%) and diarrhea (7.7%). Immune-related adverse events of any grade occurred in 23.1% of the patients with 1 (7.7%) patient experienced serious adverse events due to immune-mediated myositis. 2 patients required a dose reduction of carboplatin due to AEs but ultimately complete the prescribed treatment. There were no therapy-related death and no radiation-related dermatitis and skin hyperpigmentation.

**Table 3.**
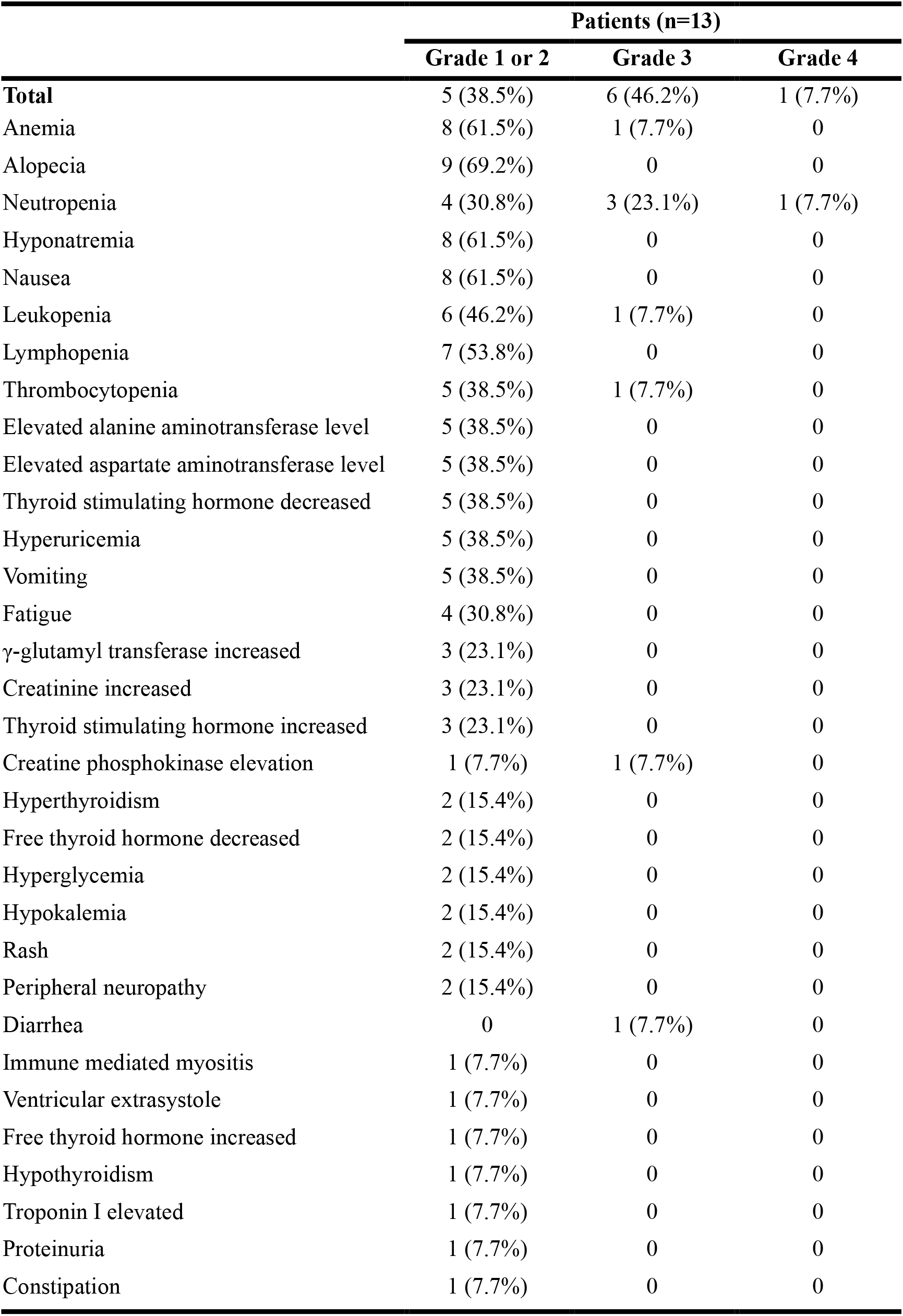
Treatment-related adverse events.

|  | Patients (n=13) |  |  |
| --- | --- | --- | --- |
|  | Grade 1 or 2 | Grade 3 | Grade 4 |
| **Total** | 5 (38.5%) | 6 (46.2%) | 1 (7.7%) |
| Anemia | 8 (61.5%) | 1 (7.7%) | 0 |
| Alopecia | 9 (69.2%) | 0 | 0 |
| Neutropenia | 4 (30.8%) | 3 (23.1%) | 1 (7.7%) |
| Hyponatremia | 8 (61.5%) | 0 | 0 |
| Nausea | 8 (61.5%) | 0 | 0 |
| Leukopenia | 6 (46.2%) | 1 (7.7%) | 0 |
| Lymphopenia | 7 (53.8%) | 0 | 0 |
| Thrombocytopenia | 5 (38.5%) | 1 (7.7%) | 0 |
| Elevated alanine aminotransferase level | 5 (38.5%) | 0 | 0 |
| Elevated aspartate aminotransferase level | 5 (38.5%) | 0 | 0 |
| Thyroid stimulating hormone decreased | 5 (38.5%) | 0 | 0 |
| Hyperuricemia | 5 (38.5%) | 0 | 0 |
| Vomiting | 5 (38.5%) | 0 | 0 |
| Fatigue | 4 (30.8%) | 0 | 0 |
| γ-glutamyl transferase increased | 3 (23.1%) | 0 | 0 |
| Creatinine increased | 3 (23.1%) | 0 | 0 |
| Thyroid stimulating hormone increased | 3 (23.1%) | 0 | 0 |
| Creatine phosphokinase elevation | 1 (7.7%) | 1 (7.7%) | 0 |
| Hyperthyroidism | 2 (15.4%) | 0 | 0 |
| Free thyroid hormone decreased | 2 (15.4%) | 0 | 0 |
| Hyperglycemia | 2 (15.4%) | 0 | 0 |
| Hypokalemia | 2 (15.4%) | 0 | 0 |
| Rash | 2 (15.4%) | 0 | 0 |
| Peripheral neuropathy | 2 (15.4%) | 0 | 0 |
| Diarrhea | 0 | 1 (7.7%) | 0 |
| Immune mediated myositis | 1 (7.7%) | 0 | 0 |
| Ventricular extrasystole | 1 (7.7%) | 0 | 0 |
| Free thyroid hormone increased | 1 (7.7%) | 0 | 0 |
| Hypothyroidism | 1 (7.7%) | 0 | 0 |
| Troponin I elevated | 1 (7.7%) | 0 | 0 |
| Proteinuria | 1 (7.7%) | 0 | 0 |
| Constipation | 1 (7.7%) | 0 | 0 |

## Discussion

This pioneering study reported the efficacy and safety of neoadjuvant SBRT in combination with adebrelimab, nab-paclitaxel and carboplatin in TNBC patients. This new therapeutic regimen achieved promising anti-tumor activity with pCR rate of 90%, and the RCB 0-I and ORR rate of 100%, respectively. More importantly, this combinatory therapeutic strategy was well tolerated in this population.

For TNBC patients who are candidates for preoperative therapy, neoadjuvant chemotherapy combined with ICIs has already achieved pCR rate of 58-64.8% in KEYNOTE-522(Schmid et al., 2020) and IMpassion031 study (Mittendorf et al., 2020). In our study, the addition of SBRT to adebrelimab and standard neoadjuvant chemotherapy achieved a significantly higher percentage of pCR (90%). Since the dose of SBRT at 24Gy in 3 fractions (bioequivalent dose [BED] = 43.2Gy) was lower than conventional preoperative radiotherapy dose (45-50Gy/23-25 fractions) (Ahmed et al., 2021), we thought that SBRT (24Gy/3f) alone could rarely achieve pCR. We speculated that the SBRT (24Gy/3f) may exert synergy with immunochemotherapy, and not just local tumor eradication effects.

Recently, SBRT has been preclinically identified as exerting immunomodulatory effects and synergizing anticancer immune responses combined with ICIs (Deng et al., 2014; Park et al., 2015). Pilones et al (Pilones et al., 2020) discovered that SBRT (two doses of 12 Gy) improved the therapeutic effects of PD-1 in a triple-negative breast cancer murine model, and this effect was enhanced by the addition of chemotherapy. TONIC trial demonstrated that SBRT (24Gy/3f) combined with nivolumab can increase the proportion of patients free of progression at 24 weeks than nivolumab alone (17% vs 8%) (Voorwerk et al., 2019). Another small sample research has indicated that preoperative SBRT (19.5-31.5Gy/3f) with neoadjuvant chemotherapy may result in fair pCR rates of 36%, with the maximum response (pCR 67%) was obtained at a dose of 25.5Gy/3f (Bondiau et al., 2013), as well as no increase in the incidences of early or late-term adverse events (AEs) (Piras et al., 2023; Takanen et al., 2022). Together, these studies suggested SBRT may have strong immunomodulatory effects, rendering a synergistic antitumor effect with immunotherapy and chemotherapy.

Another study exploring SBRT(24Gy/3f) in combination with pembrolizumab plus chemotherapy in a neoadjuvant setting for TNBC patients also achieved a promising pCR rate of 74% (PEARL) (McArthur et al., 2022). Noteworthy, PEARL study chose taxane-based chemotherapy in which 52% of patients received platinum, while our study used nab-paclitaxel and carboplatin-based regimens. It is well-established that neoadjuvant chemotherapy containing platinum has shown an increase in pCR rates of approximately 15.1% in TNBC patients (Poggio et al., 2018). Additionally, the IMpassion 130 (Emens et al., 2021) and IMpassion 131 (Miles et al., 2021) studies have suggested that nab-paclitaxel combined with ICIs may exhibit greater efficacy than paclitaxel. Thus, it is hypothesized that the selection of a chemotherapy regimen may contribute to the difference of pCR rates between the PEARL and our study (74% vs 90%). Consequently, it is imperative to determine the optimal neoadjuvant chemotherapy partners for ICIs in TNBC population.

Adverse events observed in our study were generally consistent with the known safety profiles of neoadjuvant therapy for TNBC patients in KEYNOTE-522 (Schmid et al., 2020) and IMpassion031 (Mittendorf et al., 2020) trials. The addition of SBRT did not increase any grades or grade 3 or above AEs, which were 92.3% vs 99% and 53.8% vs 57-76.8%, respectively. Furthermore, consistent with the customary toxic effects observed in lung cancers with adebrelimab plus carboplatin-based chemotherapy regimen (Wang et al., 2022; Yan et al., 2023), the addition of SBRT neither brought new AEs nor increased the incidences of grade 3 or higher or serious AEs in our population.

Our study had several limitations including a non-comparative preliminary trial with relatively small sample size, thereby impeding the comparison of our data with historical data due to insufficient statistical power. Thus, further prospective randomized clinical trial required to validate these outcomes is currently at the planning stage.

In conclusion, the addition of SBRT to adebrelimab and neoadjuvant platinum-containing therapy showed the possibility of a convenient and feasible regimen for TNBC with promising efficacy and acceptable toxicities. Neoadjuvant radiotherapy may enhance the response to immunochemotherapy through activating tumor microenvironment. Further confirmation of these findings in large-scale study is currently underway.

## Acknowledgments

Jiangsu Hengrui Pharmaceuticals Co., Ltd provided the study drug adebrelimab free of charge for patients enrolled in the study. We thank the patients and their families involved in this study.

## Additional information

### Conflict of interest

Yan Xia and Huajun Li are the employees of Jiangsu Hengrui Pharmaceuticals Co., Ltd. No other potential conflicts of interest were reported.

### Funding

This research did not receive any specific grant from funding agencies in the public, commercial, or not-for-profit sectors.

### Authors’ contributions

G. Chen, Data curation, methodology. X. Gu, Data curation, Methodology. X. Zhang, Methodology.

X. Yu, Methodology. Y. Zhang: Data curation, formal analysis, methodology. J. Xue, Methodology.

A. Li, Conceptualization, methodology. Y. Zhao, Methodology. G. He, Methodology. M. Tang, Data curation. F. Xing, Methodology. J. Yin, Methodology. X. Bian, Methodology. Y. Han, Methodology. S. Cao, Methodology. C. Liu, Formal analysis, methodology. X. Jiang, Writing-original draft. K. Zhang, Methodology. Y. Xia, Formal analysis. H. Li, Formal analysis. N. Niu, Validation, data curation, formal analysis, methodology, writing–review and editing. Caigang Liu,

Conceptualization, supervision, project administration, writing–review and editing.

### Ethics approval and consent to participate

The study was approved by Institutional Review Board of Shengjing Hospital, China Medical University and performed, according to the Declaration of Helsinki and Good Clinical Practice guidelines. Written informed consent was obtained from each patient.

### Consent for publication

All authors have read and approved the final version of this manuscript.

### Data availability statement

The raw clinical and imaging data are protected due to patient privacy laws. The datasets generated and/or analyzed during the current study are restricted from public access by related policy and law, but available from the corresponding author upon reasonable request for 10 years; de-identified clinical data and experimental data are available on request sharing, which will need the approval of the Institutional Ethical Committees. De-identified data will then be transferred to the inquiring investigator over secure file transfer.

## Notes

### Competing Interest Statement

Yan Xia and Huajun Li are the employee of Jiangsu Hengrui Pharmaceuticals Co., Ltd. No other potential conflicts of interest were reported

### Clinical Trial

Clinical trial number: NCT05132790

### Clinical Protocols

https://clinicaltrials.gov/ct2/show/NCT05132790

## References

Ahmed, M., Jozsa, F., & Douek, M. (2021). A systematic review of neo-adjuvant radiotherapy in the treatment of breast cancer. Ecancermedicalscience, 15, 1175. https://doi.org/10.3332/ecancer.2021.1175

Atun, R., Jaffray, D. A., Barton, M. B., Bray, F., Baumann, M., Vikram, B., Hanna, T. P., Knaul, F. M., Lievens, Y., Lui, T. Y., Milosevic, M., O’Sullivan, B., Rodin, D. L., Rosenblatt, E., Van Dyk, J., Yap, M. L., Zubizarreta, E., & Gospodarowicz, M. (2015, Sep). Expanding global access to radiotherapy. Lancet Oncol, 16(10), 1153–1186. https://doi.org/10.1016/S1470-2045(15)00222-3

Bondiau, P. Y., Courdi, A., Bahadoran, P., Chamorey, E., Queille-Roussel, C., Lallement, M., Birtwisle-Peyrottes, I., Chapellier, C., Pacquelet-Cheli, S., & Ferrero, J. M. (2013, Apr 1). Phase 1 clinical trial of stereotactic body radiation therapy concomitant with neoadjuvant chemotherapy for breast cancer. Int J Radiat Oncol Biol Phys, 85(5), 1193–1199. https://doi.org/10.1016/j.ijrobp.2012.10.034

Chen, Y., Gao, M., Huang, Z., Yu, J., & Meng, X. (2020, Jul 28). SBRT combined with PD-1/PD-L1 inhibitors in NSCLC treatment: a focus on the mechanisms, advances, and future challenges. J Hematol Oncol, 13(1), 105. https://doi.org/10.1186/s13045-020-00940-z

Citrin, D. E. (2017, Sep 14). Recent Developments in Radiotherapy. N Engl J Med, 377(11), 1065–1075. https://doi.org/10.1056/NEJMra1608986

Costa, R., Shah, A. N., Santa-Maria, C. A., Cruz, M. R., Mahalingam, D., Carneiro, B. A., Chae, Y. K., Cristofanilli, M., Gradishar, W. J., & Giles, F. J. (2017, Feb). Targeting Epidermal Growth Factor Receptor in triple negative breast cancer: New discoveries and practical insights for drug development. Cancer Treat Rev, 53, 111–119. https://doi.org/10.1016/j.ctrv.2016.12.010

Deng, L., Liang, H., Burnette, B., Beckett, M., Darga, T., Weichselbaum, R. R., & Fu, Y. X. (2014, Feb). Irradiation and anti-PD-L1 treatment synergistically promote antitumor immunity in mice. J Clin Invest, 124(2), 687–695. https://doi.org/10.1172/JCI67313

Emens, L. A., Adams, S., Barrios, C. H., Dieras, V., Iwata, H., Loi, S., Rugo, H. S., Schneeweiss, A., Winer, E. P., Patel, S., Henschel, V., Swat, A., Kaul, M., Molinero, L., Patel, S., Chui, S. Y., & Schmid, P. (2021, Aug). First-line atezolizumab plus nab-paclitaxel for unresectable, locally advanced, or metastatic triple-negative breast cancer: IMpassion130 final overall survival analysis. Ann Oncol, 32(8), 983–993. https://doi.org/10.1016/j.annonc.2021.05.355

Gradishar, W. J., Moran, M. S., Abraham, J., Aft, R., Agnese, D., Allison, K. H., Anderson, B., Burstein, H. J., Chew, H., Dang, C., Elias, A. D., Giordano, S. H., Goetz, M. P., Goldstein, L. J., Hurvitz, S. A., Isakoff, S. J., Jankowitz, R. C., Javid, S. H., Krishnamurthy, J., Leitch, M., Lyons, J., Mortimer, J., Patel, S. A., Pierce, L. J., Rosenberger, L. H., Rugo, H. S., Sitapati, A., Smith, K. L., Smith, M. L., Soliman, H., Stringer-Reasor, E. M., Telli, M. L., Ward, J. H., Wisinski, K. B., Young, J. S., Burns, J., & Kumar, R. (2022, Jun). Breast Cancer, Version 3.2022, NCCN Clinical Practice Guidelines in Oncology. J Natl Compr Canc Netw, 20(6), 691–722. https://doi.org/10.6004/jnccn.2022.0030

Han, M. G., Wee, C. W., Kang, M. H., Kim, M. J., Jeon, S. H., & Kim, I. A. (2022, May 29). Combination of OX40 Co-Stimulation, Radiotherapy, and PD-1 Inhibition in a Syngeneic Murine Triple-Negative Breast Cancer Model. Cancers (Basel), 14(11). https://doi.org/10.3390/cancers14112692

Holyoake, D. L. P., Robinson, M., Silva, M., Grose, D., McIntosh, D., Sebag-Montefiore, D., Radhakrishna, G., Mukherjee, S., & Hawkins, M. A. (2021, Feb). SPARC, a phase-I trial of pre-operative, margin intensified, stereotactic body radiation therapy for pancreatic cancer. Radiother Oncol, 155, 278–284. https://doi.org/10.1016/j.radonc.2020.11.007

Keenan, T. E., & Tolaney, S. M. (2020, Apr). Role of Immunotherapy in Triple-Negative Breast Cancer. J Natl Compr Canc Netw, 18(4), 479–489. https://doi.org/10.6004/jnccn.2020.7554

Kishi, N., Kanayama, N., Hirata, T., Ohira, S., Wada, K., Kawaguchi, Y., Konishi, K., Nagata, S., Nakatsuka, S. I., Marubashi, S., Tomokuni, A., Wada, H., Kobayashi, S., Tomita, Y., & Teshima, T. (2020, Mar 5). Preoperative Stereotactic Body Radiotherapy to Portal Vein Tumour Thrombus in Hepatocellular Carcinoma: Clinical and Pathological Analysis. Sci Rep, 10(1), 4105. https://doi.org/10.1038/s41598-020-60871-0

Liu, Y., Veale, C., Hablitz, D., Krontiras, H., Dalton, A., Meyers, K., Dobelbower, M., Lancaster, R., Bredel, M., Parker, C., Keene, K., Thomas, E., & Boggs, D. (2022). Feasibility and Short-Term Toxicity of a Consecutively Delivered Five Fraction Stereotactic Body Radiation Therapy Regimen in Early-Stage Breast Cancer Patients Receiving Partial Breast Irradiation. Front Oncol, 12, 901312. https://doi.org/10.3389/fonc.2022.901312

Loi, S., Drubay, D., Adams, S., Pruneri, G., Francis, P. A., Lacroix-Triki, M., Joensuu, H., Dieci, M. V., Badve, S., Demaria, S., Gray, R., Munzone, E., Lemonnier, J., Sotiriou, C., Piccart, M. J., Kellokumpu-Lehtinen, P. L., Vingiani, A., Gray, K., Andre, F., Denkert, C., Salgado, R., & Michiels, S. (2019, Mar 1). Tumor-Infiltrating Lymphocytes and Prognosis: A Pooled Individual Patient Analysis of Early-Stage Triple-Negative Breast Cancers. J Clin Oncol, 37(7), 559–569. https://doi.org/10.1200/JCO.18.01010

McArthur, H. L., Shiao, S., Karlan, S., Basho, R., Amersi, F., Burnison, M., Mirhadi, A., Chung, A., Chung, C. T., Dang, C., Richardson, H., Giuliano, A. E., Kapoor, N., Larkin, B., Godinez, H., Dunn, S. A., Khameneh, N. H., Knott, S., McAndrew, P., Mita, M., Park, D. J., Abaya, C., Chen, J. H., Ly, A., Bossuyt, V., & Ho, A. (2022). Abstract PD10-01: The PEARL trial: Pre-operative pembrolizumab with radiation therapy in early stage triple negative breast cancer. Cancer Research, 82(4_Supplement), PD10-01–PD10-01. https://doi.org/10.1158/1538-7445.Sabcs21-pd10-01

Miles, D., Gligorov, J., Andre, F., Cameron, D., Schneeweiss, A., Barrios, C., Xu, B., Wardley, A., Kaen, D., Andrade, L., Semiglazov, V., Reinisch, M., Patel, S., Patre, M., Morales, L., Patel, S. L., Kaul, M., Barata, T., O’Shaughnessy, J., & investigators, I. M. (2021, Aug). Primary results from IMpassion131, a double-blind, placebo-controlled, randomised phase III trial of first-line paclitaxel with or without atezolizumab for unresectable locally advanced/metastatic triple-negative breast cancer. Ann Oncol, 32(8), 994–1004. https://doi.org/10.1016/j.annonc.2021.05.801

Mittendorf, E. A., Zhang, H., Barrios, C. H., Saji, S., Jung, K. H., Hegg, R., Koehler, A., Sohn, J., Iwata, H., Telli, M. L., Ferrario, C., Punie, K., Penault-Llorca, F., Patel, S., Duc, A. N., Liste-Hermoso, M., Maiya, V., Molinero, L., Chui, S. Y., & Harbeck, N. (2020, Oct 10). Neoadjuvant atezolizumab in combination with sequential nab-paclitaxel and anthracycline-based chemotherapy versus placebo and chemotherapy in patients with early-stage triple-negative breast cancer (IMpassion031): a randomised, double-blind, phase 3 trial. Lancet, 396(10257), 1090–1100. https://doi.org/10.1016/S0140-6736(20)31953-X

Nicosia, L., Figlia, V., Ricottone, N., Cuccia, F., Mazzola, R., Giaj-Levra, N., Ricchetti, F., Rigo, M., Jafari, F., Maria Magrini, S., Girlando, A., & Alongi, F. (2022, Aug). Stereotactic body radiotherapy (SBRT) and concomitant systemic therapy in oligoprogressive breast cancer patients. Clin Exp Metastasis, 39(4), 581–588. https://doi.org/10.1007/s10585-022-10167-6

Novikov, S. N., Gafton, G. I., Ebert, M. A., Fedosova, E. A., Melnik, J. S., Zinovev, G. V., Gafton, I. G., Sinyachkin, M. S., & Kanaev, S. V. (2021, Aug). Preoperative stereotactic ablative body radiotherapy with postoperative conventional irradiation of soft tissue sarcomas: Protocol overview with a preliminary safety report. Radiother Oncol, 161, 126–131. https://doi.org/10.1016/j.radonc.2021.05.025

Park, S. S., Dong, H., Liu, X., Harrington, S. M., Krco, C. J., Grams, M. P., Mansfield, A. S., Furutani, K. M., Olivier, K. R., & Kwon, E. D. (2015, Jun). PD-1 Restrains Radiotherapy-Induced Abscopal Effect. Cancer Immunol Res, 3(6), 610–619. https://doi.org/10.1158/2326-6066.CIR-14-0138

Pilones, K. A., Hensler, M., Daviaud, C., Kraynak, J., Fucikova, J., Galluzzi, L., Demaria, S., & Formenti, S. C. (2020, Oct 20). Converging focal radiation and immunotherapy in a preclinical model of triple negative breast cancer: contribution of VISTA blockade. Oncoimmunology, 9(1), 1830524. https://doi.org/10.1080/2162402X.2020.1830524

Piras, A., Sanfratello, A., Boldrini, L., D’Aviero, A., Pernice, G., Sortino, G., Valerio, M. R., Gennari, R., D’Angelo, I., Marazzi, F., Angileri, T., & Daidone, A. (2023). Stereotactic Radiotherapy in Early-Stage Breast Cancer in Neoadjuvant and Exclusive Settings: A Systematic Review. Oncol Res Treat, 46(3), 116–123. https://doi.org/10.1159/000528640

Poggio, F., Bruzzone, M., Ceppi, M., Ponde, N. F., La Valle, G., Del Mastro, L., de Azambuja, E., & Lambertini, M. (2018, Jul 1). Platinum-based neoadjuvant chemotherapy in triple-negative breast cancer: a systematic review and meta-analysis. Ann Oncol, 29(7), 1497–1508. https://doi.org/10.1093/annonc/mdy127

Schmid, P., Cortes, J., Pusztai, L., McArthur, H., Kummel, S., Bergh, J., Denkert, C., Park, Y. H., Hui, R., Harbeck, N., Takahashi, M., Foukakis, T., Fasching, P. A., Cardoso, F., Untch, M., Jia, L., Karantza, V., Zhao, J., Aktan, G., Dent, R., O’Shaughnessy, J., & Investigators, K.-. (2020, Feb 27). Pembrolizumab for Early Triple-Negative Breast Cancer. N Engl J Med, 382(9), 810–821. https://doi.org/10.1056/NEJMoa1910549

Takanen, S., Pinnaro, P., Farina, I., Sperati, F., Botti, C., Vici, P., Soriani, A., Marucci, L., & Sanguineti, G. (2022). Stereotactic partial breast irradiation in primary breast cancer: A comprehensive review of the current status and future directions. Front Oncol, 12, 953810. https://doi.org/10.3389/fonc.2022.953810

Viani, G. A., Gouveia, A. G., Louie, A. V., Korzeniowski, M., Pavoni, J. F., Hamamura, A. C., & Moraes, F. Y. (2021, Nov). Stereotactic body radiotherapy to treat breast cancer oligometastases: A systematic review with meta-analysis. Radiother Oncol, 164, 245–250. https://doi.org/10.1016/j.radonc.2021.09.031

Voorwerk, L., Slagter, M., Horlings, H. M., Sikorska, K., van de Vijver, K. K., de Maaker, M., Nederlof, I., Kluin, R. J. C., Warren, S., Ong, S., Wiersma, T. G., Russell, N. S., Lalezari, F., Schouten, P. C., Bakker, N. A. M., Ketelaars, S. L. C., Peters, D., Lange, C. A. H., van Werkhoven, E., van Tinteren, H., Mandjes, I. A. M., Kemper, I., Onderwater, S., Chalabi, M., Wilgenhof, S., Haanen, J., Salgado, R., de Visser, K. E., Sonke, G. S., Wessels, L. F. A., Linn, S. C., Schumacher, T. N., Blank, C. U., & Kok, M. (2019, Jun). Immune induction strategies in metastatic triple-negative breast cancer to enhance the sensitivity to PD-1 blockade: the TONIC trial. Nat Med, 25(6), 920–928. https://doi.org/10.1038/s41591-019-0432-4

Wang, J., Zhou, C., Yao, W., Wang, Q., Min, X., Chen, G., Xu, X., Li, X., Xu, F., Fang, Y., Yang, R., Yu, G., Gong, Y., Zhao, J., Fan, Y., Liu, Q., Cao, L., Yao, Y., Liu, Y., Li, X., Wu, J., He, Z., Lu, K., Jiang, L., Hu, C., Zhao, W., Zhang, B., Shi, W., Zhang, X., Cheng, Y., & Group, C.-S. (2022, Jun). Adebrelimab or placebo plus carboplatin and etoposide as first-line treatment for extensive-stage small-cell lung cancer (CAPSTONE-1): a multicentre, randomised, double-blind, placebo-controlled, phase 3 trial. Lancet Oncol, 23(6), 739–747. https://doi.org/10.1016/S1470-2045(22)00224-8

Yan, W., Zhong, W. Z., Liu, Y. H., Chen, Q., Xing, W., Zhang, Q., Liu, L., Ge, D., Chen, K., Yang, F., Lin, X., Song, L., Shi, W., & Wu, Y. L. (2023, Feb). Adebrelimab (SHR-1316) in Combination With Chemotherapy as Perioperative Treatment in Patients With Resectable Stage II to III NSCLCs: An Open-Label, Multicenter, Phase 1b Trial. J Thorac Oncol, 18(2), 194–203. https://doi.org/10.1016/j.jtho.2022.09.222

